# AI as a signal assessor – Can a Large Language Model perform causality assessment on a case series?

**DOI:** 10.64898/2026.06.26.26356656

**Authors:** Aditi Shenoy, Alem Zekarias, Anders Viklund, Joseph Mitchell, Jim Barrett, Lovisa Sandberg, Eva-Lisa Meldau, Henric Taavola-Gustafsson

## Abstract

**Background:** Large Language Models (LLMs) are increasingly explored for pharmacovigilance tasks, including information extraction, case documentation, and single-case causality assessment. However, their ability to support causality assessment at the case series level — a complex, time-intensive task requiring clinical reasoning across multiple reports — remains unexplored.

**Objective:** To investigate how a large-scale general-purpose LLM can support pharmacovigilance professionals in assessing causality in a case series, and to explore how prompt design influences the quality of the model’s reasoning.

**Methods:** GPT-4o was used to assess causality for five drug – adverse event combinations, using an adaptation of the Bradford Hill viewpoints for case series assessment. The combinations represented varying drugs and vaccines, adverse events, and case series sizes (5–402 reports). One combination served as a negative control. Structured prompts were iteratively developed and refined using one combination, then applied to all combinations. LLM-generated assessments for each viewpoint were qualitatively evaluated by human annotators for accuracy (precision), and the LLM’s coverage of key aspects from the original signal text was assessed for one combination (recall).

**Results:** Across all five combinations, annotators agreed with 79–92% of the LLM’s output sentences. Full disagreement was consistently low (3–7%), with errors typically involving misinterpretation of complex report details rather than outright fabrication. Prompt design substantially influenced output quality; providing Bradford Hill viewpoint descriptions, including case series data, and adding explicit anti-hallucination instructions improved specificity and grounding. For the recall assessment, 15 of 23 key segments from the original signal text were reflected in the LLM output. The overall summary assessments demonstrated balanced reasoning, correctly distinguishing between positive safety signals and the negative control, and provided a coherent synthesis suitable as a starting point for human assessors.

**Conclusions:** LLMs have the potential to generate contextually nuanced and largely accurate preliminary causality assessments of case series aligned with the Bradford Hill viewpoints, with a low but non-zero hallucination rate. These findings support LLMs as a tool to augment, not replace, expert judgment in signal assessment. Future work should address larger and more diverse signal sets, improved evaluation frameworks for generative output, and the integration of pre-computed summary statistics to reduce errors.

**Key points:**

- A large language model (GPT-4o) can support pharmacovigilance professionals by generating preliminary causality assessments of a case series that align with human expert judgment.
- Generated in minutes, such assessments could give assessors a structured starting point for in-depth review, condensing case-level information and surfacing early evidence for or against a signal.
- Output quality depended heavily on prompt design, and because the hallucination rate was low but not zero, these tools should augment, not replace, expert judgment in signal assessment.

## 1. Introduction

Large Language Models (LLMs) have made substantial advances in medicine, especially regarding information extraction, literature screening, summarizing, paraphrasing, and translating clinical text [1]. Recently, an increasing number of studies have shown the effectiveness of generative artificial intelligence (GenAI) and LLMs in pharmacovigilance (PV) activities [2]. LLMs are being explored for use in PV processes, such as case documentation [3], information extraction [4,5], detecting clinically significant drug-drug interactions that may lead to an adverse event [6], and causality assessment for single cases [7].

Additionally, the increasing volume of adverse event reports reflects growing engagement and provides valuable opportunities for improving drug safety. To fully leverage this source of information, more capacity and streamlined processes for efficient review are needed. This is especially true for signal assessment, being a time-intensive process that demands careful evaluation of the causal link between the adverse event and a specific drug or vaccine. Signal assessment is not harmonized across the field and is approached in many different ways, with the rigor and structure of causal evaluation varying between assessors and organizations. The LLM’s ability to parse and comprehend large amounts of text could assist assessors in this process and potentially accelerate the identification of safety signals.

One structured approach that can support such causal reasoning is the Bradford Hill (BH) viewpoints [8], originally developed for epidemiological analysis in the context of occupational medicine. While not designed for pharmacovigilance, the viewpoints can serve as inspiration for structuring causality assessment of case series. These viewpoints comprise nine key considerations for assessors — *strength*, *consistency*, *specificity*, *temporality*, *biological gradient*, *plausibility*, *coherence*, *experimental evidence*, and *analogy*. These are especially valuable as they provide a structured approach to guide causal reasoning when working with incomplete or missing data, which is often the case with adverse event reports.

For the scope of this study we have focused on BH since it is a widely used framework for causality assessment in epidemiologic studies [9] and we have used an adaptation for case series assessment in PV based on the work of Shakir and Layton [10] that we further adapted for use with an LLM. Other well-known approaches to causality assessment, such as the WHO-UMC causality assessment system [11] and Naranjo criteria [12], are intended for single-case assessment rather than case series, and were therefore not applicable to this study.

The number of reports in a case series varies significantly and for a PV assessor to evaluate an entire case series of reports for a drug – adverse event combination and evaluate whether it is plausible or not is a non-trivial and often time-consuming process. Hence, LLMs could play a role in summarizing the case series, creating an initial assessment and providing an overall summary of the evidence, or lack thereof, for causality, to serve as a starting point for the human assessor. We have used an LLM to estimate its performance on causality assessment for a few case series of signals with varying characteristics (e.g. number and quality of reports in the case series, drug involved, type of adverse event).

Despite the growing literature on the use of GenAI in PV practice, there is limited evidence on how well LLMs can reason clinically. Clinical reasoning is needed for all the BH viewpoints, but especially for viewpoints such as *plausibility* and *coherence*, which rely heavily on the PV assessor’s medical and pharmacological knowledge. Since LLMs have been trained on large corpora of text that include medical knowledge, they may have the capacity to draw on this knowledge when performing such assessments.

Given the rapid evolution of Artificial Intelligence (AI) technologies, we intentionally designed this study as an exploratory, preliminary investigation. Through this work, we examine how well LLMs are able to reason around causality assessment using a series of reports of adverse events of medicines and vaccines (hereafter ‘drugs’), and explore how prompt-design choices may influence this reasoning. Our goal was to generate initial findings, using pragmatic design choices, to stimulate discussion and encourage more comprehensive and rigorous future research in this area.

The research question for this study is; How can a large-scale general-purpose LLM support PV professionals in assessing causality in a case series, particularly through the lens of the BH viewpoints?

## 2. Methods

### 2.1. Model selection

Given the promise generative pre-trained transformer (GPT) models have shown with clinical reasoning and natural language processing (NLP) tasks with medical data [13,14], an evaluation of their capability for causal reasoning in PV signal assessment was performed. To this end, OpenAI’s GPT-4o was chosen for the experiments. This model should have inherent medical knowledge from its training data and a large enough context window (amount of text it can consider at once) to be able to use a meaningful amount of the case series data and generate assessments for each of the BH viewpoints.

To limit the randomness of the output, we set the model parameter “temperature” to 0. Typical values for the “temperature” are considered to be between 0 and 1 and can be seen as a way to control how creative and random the model output will be. When set to 0, the output tends to produce similar output every time it gets the same input, while setting it to 1 will produce a more varied output for the same input. [15]

GPT-4o represents a state-of-the-art LLM model that was also generally accessible^1^ at the time when this study was conducted, between June and August 2025. It has good reasoning performance without being a dedicated reasoning model. Reasoning models like OpenAI’s o3 and o4-mini are trained to ‘think’ before they generate a response by producing text representing the “thinking steps” it needs to take before generating the final answer [16]. Since access to these reasoning models through a secure and private manner was limited, they were not used in this study.

### 2.2. Data privacy considerations

All data within the used reports were checked to ensure they did not contain any sensitive information. Further steps were taken to ensure that the data sent to the models for processing were not used by Microsoft or OpenAI for training future versions of the models, nor retained for any purpose. All AI processing was performed using Azure OpenAI API Service within the Microsoft’s Azure cloud platform. Data remained in Azure while processing and was not accessible by Microsoft staff, nor shared with OpenAI or used for model training. For enhanced privacy, a configuration that disables even temporary storage of inputs and outputs was used, so no data was retained beyond immediate processing. Microsoft’s security and privacy standards were verified to comply with Uppsala Monitoring Centre’s internal policies.

### 2.3. Dataset selection and preprocessing

To evaluate the causality assessment capacity of the LLM, safety signals of adverse events of medicinal products by Uppsala Monitoring Centre were selected. These drug – adverse event combinations were *omeprazole – cutaneous vasculitis*, *covid-19 vaccine – vulval ulceration* [17], *methotrexate – muscle spasms* [18], and *dolutegravir – sexual dysfunction* [19]. These were selected to represent a wide variety of drugs, adverse events, and number of reports in the case series. Additionally, we included a combination, *pazopanib – back pain*, to serve as a negative control, for which the assessment did not find evidence of a causal association. See Table 1 for a list and additional details on all the included combinations.

**Table 1:**
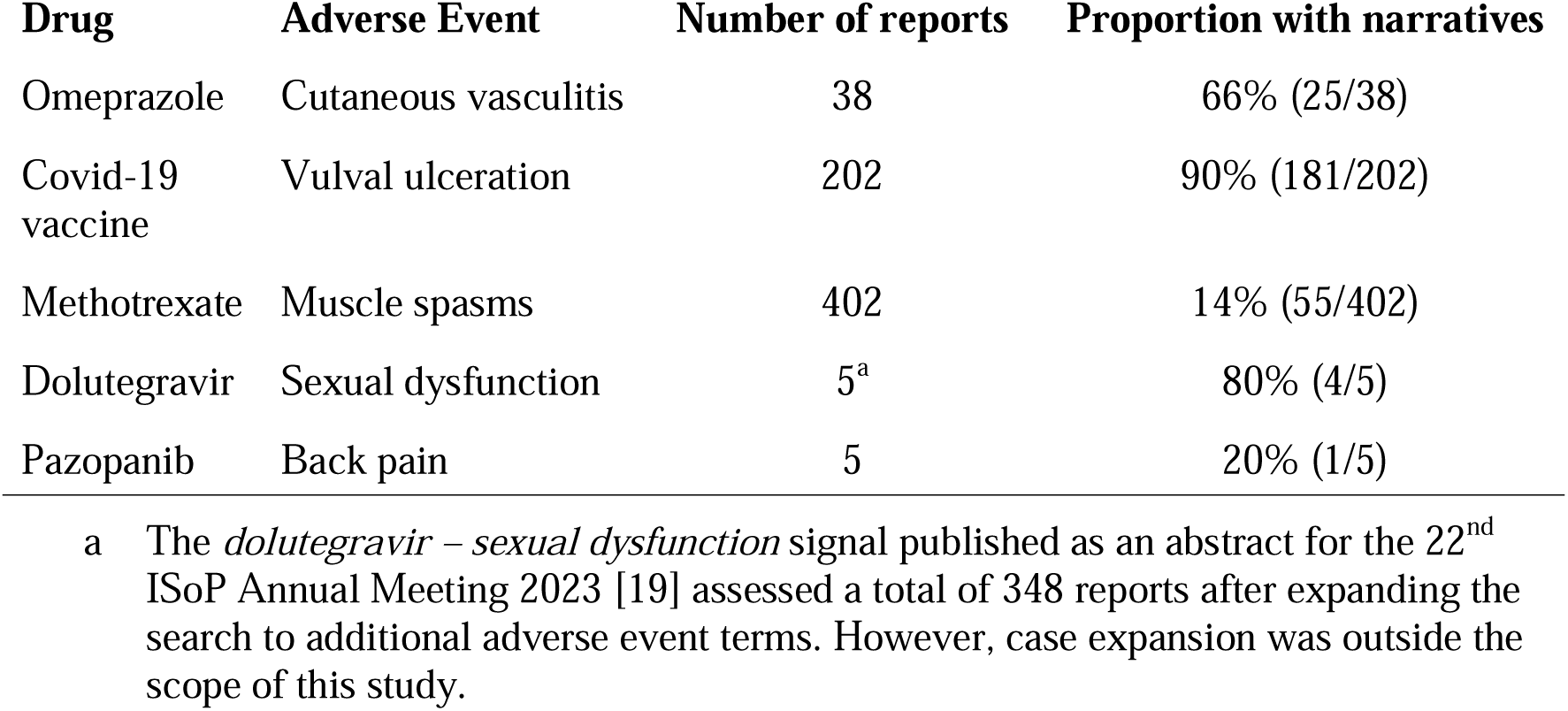
Drug – adverse event combinations selected for evaluating causality assessment by Large Language Models (LLMs)

| Drug | Adverse Event | Number of reports | Proportion with narratives |
| --- | --- | --- | --- |
| Omeprazole | Cutaneous vasculitis | 38 | 66% (25/38) |
| Covid-19 vaccine | Vulval ulceration | 202 | 90% (181/202) |
| Methotrexate | Muscle spasms | 402 | 14% (55/402) |
| Dolutegravir | Sexual dysfunction | 5<sup>a</sup> | 80% (4/5) |
| Pazopanib | Back pain | 5 | 20% (1/5) |

<sup>a</sup> The *dolutegravir – sexual dysfunction* signal published as an abstract for the 22<sup>nd</sup> ISoP Annual Meeting 2023 [19] assessed a total of 348 reports after expanding the search to additional adverse event terms. However, case expansion was outside the scope of this study.

As input data for the LLM, reports from VigiBase [20], the WHO global database of adverse event reports for medicines and vaccines, was used. To ensure that the input data for the LLM closely matched the information available to assessors during the original signal evaluation, we included only those reports for each drug – adverse event combination that were available at the time of the initial assessment. Reports received or updated after the signal was assessed, as well as suspected duplicates (identified by vigiMatch [21]), were excluded. Excluding updated reports was a deliberately conservative choice: we wanted to avoid giving the LLM any advantage over the original human assessors, and accepted that it could instead place the LLM at a disadvantage, since updated reports — much like additional reports — could contain information not available at the time of the original assessment. As a result, the LLM had access to fewer cases than the human assessors (e.g., for *omeprazole – cutaneous vasculitis*, 38 instead of 47 reports).

The signal for *omeprazole – cutaneous vasculitis* was used to develop and refine prompts for the LLM. The remaining four combinations were evaluated on the responses generated using this refined prompt.

For each report, a generated unique report ID number, source country of the report, patient sex, patient age, seriousness of the report (yes/no), dosage of the drug of interest, time-to-onset (TTO) of the event of interest, date of last update of the report, and the free-text case narrative describing the reported clinical course and circumstances were extracted. For every reported event, the MedDRA^®^ preferred term name^2^ [22], event start and end dates, and reported outcome were made available. For all drugs marked as suspected or interacting within each report, the active ingredient name, start and end dates, indication, action taken (e.g. drug withdrawn or dose changed), and administration route were extracted. Additionally, for reported medical history, the medical term, start and end dates, and continuation status (yes/no) were extracted.

For each drug – adverse event combination, the reports were fed to the LLM in descending order of each report’s vigiGrade completeness score [23], so that when not all reports could be processed, the most complete reports were prioritized. The score is based on the completeness of the information contained within each report. There is a score for each combination of drug and adverse event on the report as well as an overall score for the full report. Since a single report can have multiple entries for the same drug – adverse event combination, the highest score for the combination of interest was used.

For the drug – adverse event combinations with very large case series (exceeding 100 reports), some filtering criteria were applied due to the token (word or part of word) limit imposed by the LLM models. The aim of the filtering was to select the most relevant reports for the LLM to use. If a response was not generated due to hitting the maximum token limit, another attempt was made using only the reports containing narratives. For the combination *Covid-19 vaccine – vulval ulceration*, the above filtering criteria yielded 53 case reports which were able to be processed. For the combination *methotrexate – muscle spasms*, the above filtering criteria resulted in 195 reports, which still exceeded the LLM maximum token limit. So, one additional criterion of vigiGrade completeness score ≥ 0.8 yielded 18 case reports that fit within the token usage limit.

### 2.4. Experimental design

#### 2.4.1. Prompt design

The impact of prompt design on assessments of the BH viewpoints was evaluated. This included making changes to both the system and user prompts. The system prompt outlines the model’s role and defines constraints such as instructions to avoid hallucinations or making up citations. The user prompt provides the context, task, case series and output format instructions. Variations in prompt structure, such as inclusion of headers, verbose instructions for clarity, as well as the effect of additional data and ways to reduce hallucinations, were studied.

There were two main prompts (both consisting of a system and user prompt) used in this study, one template prompt for running the assessment for each of the BH viewpoints, where the main difference was the provided description specified for each viewpoint, and one prompt that used the assessments generated for each viewpoint from the previous prompt to produce an overall summary assessment for each drug – adverse event combination.

For the BH viewpoints, an adaptation for case series assessment based on [10] was further refined for use by the LLM. All viewpoint specifications can be found in the electronic supplementary material Section 1. For the BH *strength* viewpoint, we utilized disproportionality analysis metrics, including the Information Component (IC) and Reporting Odds Ratio (ROR), as these are commonly used quantitative measures in PV for evaluating the magnitude of association between a drug and an adverse event. However, it is important to note that disproportionality analysis effect sizes do not reliably reflect the strength of association in the same way as in pharmacoepidemiological studies. Disproportionality analysis metrics are subject to confounding factors such as reporting biases, differences in exposure, and lack of denominator data, and therefore should not be considered a replacement or proxy for epidemiological effect size. While disproportionality analysis was used to inform the *strength* viewpoint in this study, we do not recommend its use as a substitute for effect size in causal inference. Disproportionality analysis was only included in this study to investigate how the LLM would reason around it. In addition to disproportionality analysis, the description of the *strength* viewpoint provided to the LLM also included positive de- and rechallenge as indicators of strength. This deviates from both the original BH *strength* viewpoint [8] and the Shakir and Layton adaptation [10] on which our implementation is based. In our descriptions, dechallenge and rechallenge are not just important for the *strength* viewpoint but even considered an important element of the *temporality* viewpoint, both providing relevant timeline information.

#### 2.4.2. Qualitative evaluation of Bradford Hill viewpoints

To evaluate the LLM’s capability to support signal assessment of a case series using the BH viewpoints, both the accuracy of its output statements and the extent to which it included key aspects from the original signal text were qualitatively examined.

For this, each of the LLM responses for the nine BH viewpoints across the five drug – adverse event combinations were split into sentences. These were reviewed by four of the authors of this article as annotators: two pharmacists with MSc degrees and pharmacovigilance experience (AZ with 13 years; AV with 10 years) and two data scientists with PhDs and pharmacovigilance experience (AS with 2 years; HTG with 10 years). For *omeprazole – cutaneous vasculitis,* all the LLM sentences were reviewed, while for the other combinations a target was set to review 50% of sentences within the available timeframe allotted to the study. Each sentence was compared to the information available in the case series data that was available to the LLM and general pharmacological knowledge (combined with searching the literature when needed) to decide whether the annotators agreed with the statement, answering “yes”, “no” or “partial”. This can thus be seen as a precision test with manual annotation to evaluate the degree of hallucinations from the LLM.

To investigate to what extent the LLM was able to capture all the relevant information to make an adequate assessment of causality, the *omeprazole – cutaneous vasculitis* signal text was used as the ground truth. Key segments of the signal text were identified and mapped to their corresponding BH viewpoint. This enabled a determination of whether any critical aspects of the signal assessment had been overlooked by the LLM. One annotator (AV) conducted a manual review to determine whether these identified key elements were reflected in the BH summaries generated by the LLM. This approach effectively served as a recall assessment using manual annotation.

#### 2.4.3. Manual review of overall assessments

The overall summarized assessments of the combinations made by the LLM based on the intermediate individual LLM assessments per BH viewpoint were manually reviewed for accuracy. This was done for two of the combinations, *omeprazole – cutaneous vasculitis* and the negative control, *pazopanib – back pain*.

## 3. Results

### 3.1. Prompt design

In contrast to traditional machine learning methods which are trained using annotated data, LLMs are instructed to perform tasks using so-called prompts, carefully designed plain text instructions. This makes prompt design an important step in developing a model to perform the task at hand. This includes making changes to both the system prompt – which outlines the model’s role and defines constraints to avoid hallucinations or making up citations in the model’s responses – and the user prompt – where the context, task, case series, and output format instruction are provided. While LLMs possess inherent biomedical knowledge, their performance for clinical reasoning is notably enhanced when structured, context-rich prompts are used, such as the inclusion of headers, or verbose instructions for clarity. The final prompt we designed, which was used for all subsequent experiments, can be found in the electronic supplementary material Table S1.

The following additions to the prompt proved valuable during prompt design:

1. Providing descriptions of the BH viewpoints made model responses more specific and ensured reflections around the viewpoints.
2. Adding the disclaimer, ‘You should clearly distinguish between well-supported mechanisms and speculative or incorrect ones. Do not make up references or hallucinate molecular interactions. Do not use epidemiological or case-level data unless explicitly provided.’ to the end of the system prompt allowed the model responses to be restricted to information provided in the case series and not refer to such information it might have seen in its training data. For details see the electronic supplementary material Table S2.
3. Including case series data is important to make the assessment specific and anchored in evidence from the case series, as compared to solely relying on the LLM’s general knowledge about pharmacology, pathophysiology, and drug mechanisms. Exceptions to this are *plausibility* and *coherence*, which apply to the drug – adverse event combination level and case data confuses the model with unnecessary detail. See section 3.2 below.
4. The type of information provided for the case series is of critical importance. Inclusion of TTO and dechallenge/rechallenge data provide clear and relevant timelines for the *temporality* and *strength* viewpoints, providing additional information about whether the adverse event occurred after drug administration and whether the timing aligns with pharmacological expectations. Rechallenge data, as seen in some Covid-19 vaccine reports, further supports causality by demonstrating recurrence upon re-exposure, which helps establish *temporality* by reinforcing the order of drug exposure and adverse event. Nevertheless, these assessments can be subject to overconfidence and may analyze the viewpoints in isolation, disregarding for example alternative explanations.

### 3.2. Qualitative evaluation of Bradford Hill viewpoints

#### 3.2.1. Hallucination assessment

The proportion of annotated sentences was 100% for *omeprazole – cutaneous vasculitis* and for the remaining four drug – adverse event combinations the proportion ranged from 37% to 67%. See Figure 1 (left).

**Fig. 1.**
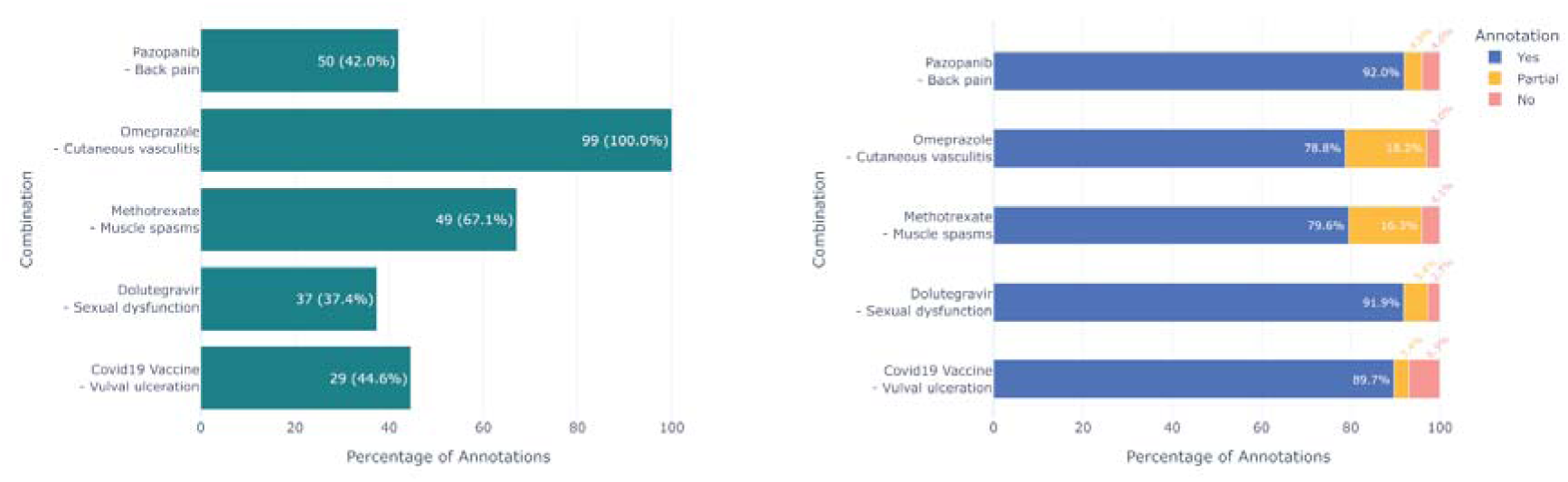
(left) Proportion of annotated sentences per drug – adverse event combination. (right) Proportion of annotated sentences where the annotator agreed (yes), partially agreed (partial), and did not agree (no) with the output of the LLM.

For the majority of the annotated sentences (79% - 92%, depending on drug – adverse event combination), annotators agreed with the LLM output. Sentences considered to contain hallucinations, defined as those with partial agreement or no agreement with the annotators, accounted for 3% - 18% and 3% - 7%, respectively. These proportions were consistently low for the sentences with no agreement across the five drug – adverse event combinations. See Figure 1 (right).

When looking at the sentences where the annotator did not agree with the LLM, the errors were not generally outright wrong, but rather confusions. For example, it considered there being a positive de-challenge for the drug of interest (methotrexate), when it was actually positive for methotrexate and another drug being discontinued at the same time. The report then describes how methotrexate was re-introduced without re-occurrence of the adverse event, but the LLM failed to take this into account. Another example is when the LLM was confused by a negative calculated TTO on a report concerning covid-19 vaccine – vulval ulceration, where the negative TTO was referring to the second dose of the vaccine that was also present on the report, but the adverse event happened after the first dose which then had a positive TTO. There were also statements of there being a lack of reports for similar drugs, which is only incorrect due to the LLM only being given the reports for the drug – adverse event combination of interest and no ability to search for other reports, which a human assessor would have.

For the sentences where the annotator agreed with the LLM, we see successful reasoning regarding pharmacological knowledge and complex reports. For example, it was able to draw a parallel between omeprazole and cutaneous vasculitis where it stated that cutaneous vasculitis can be immune-mediated and that proton pump inhibitors have been known to cause hypersensitivity reactions. Another example is where it recognized that a report where the patient had multiple medications would complicate the assessment of causality. See the LLM output examples in Table 2.

**Table 2.**
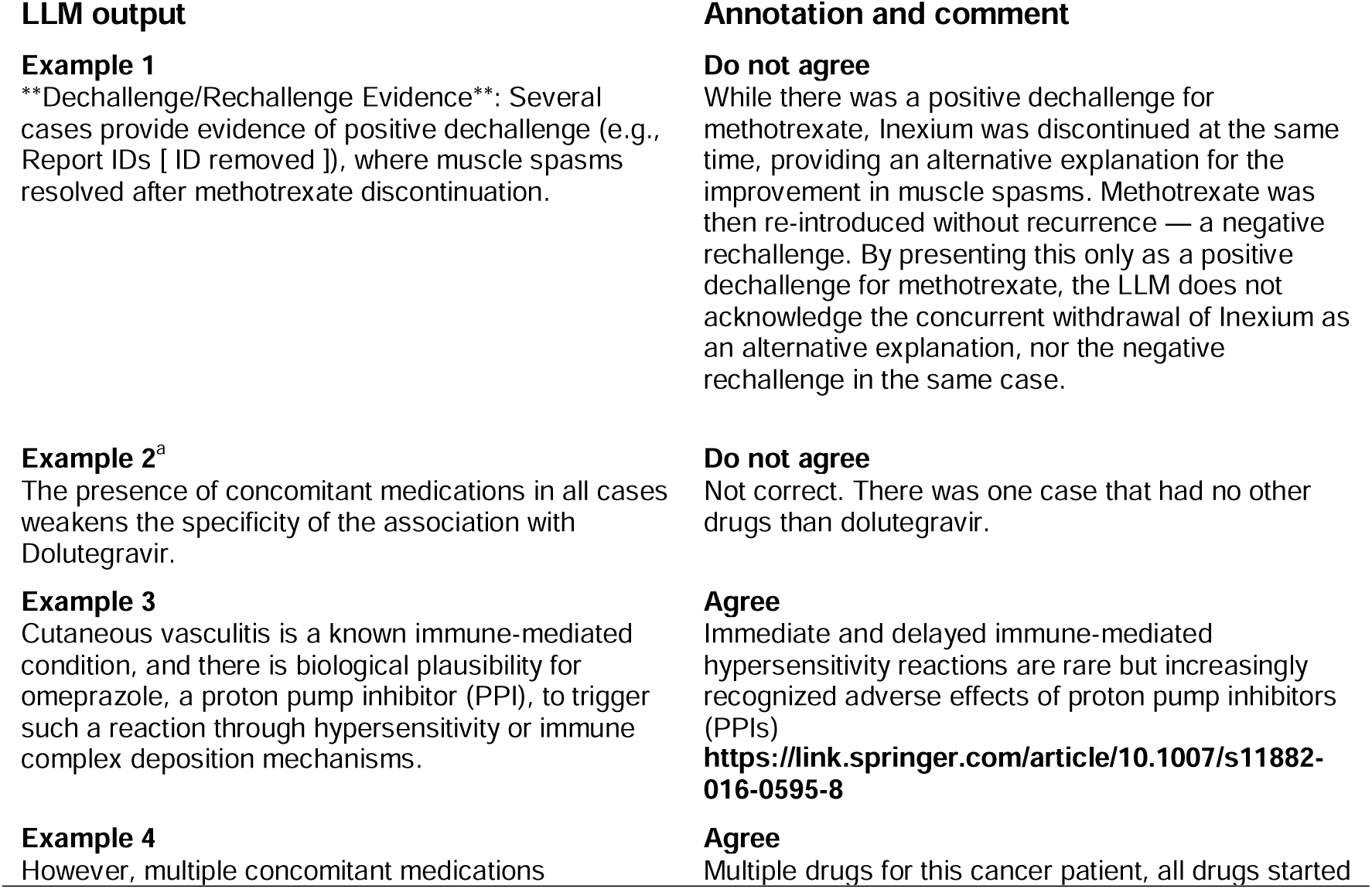
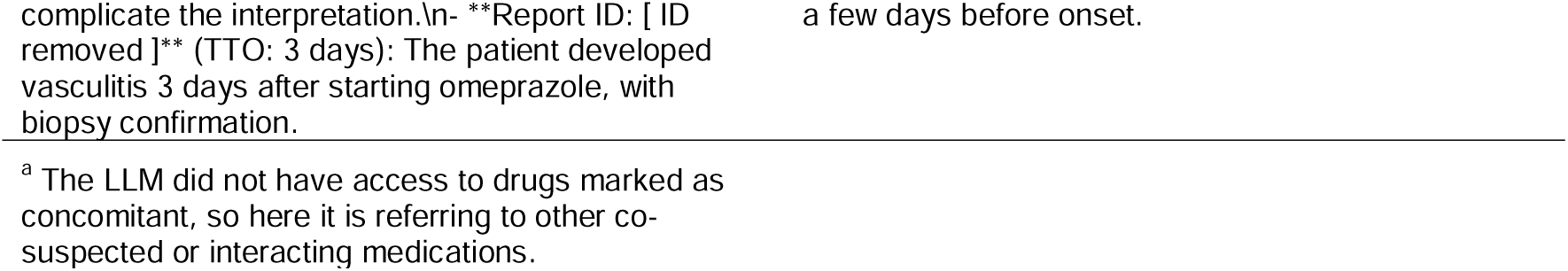
Examples of LLM output along with annotation and comment. Annotations were made conservatively: where the LLM’s reasoning was partially correct but could be misleading or omitted relevant nuance, it was rated as a disagreement rather than agreement

When aggregating the data per BH viewpoint the proportion of annotated sentences ranges from 25% for *temporality* to 90% for *biological gradient*. See Figure 2 (left).

**Fig. 2.**
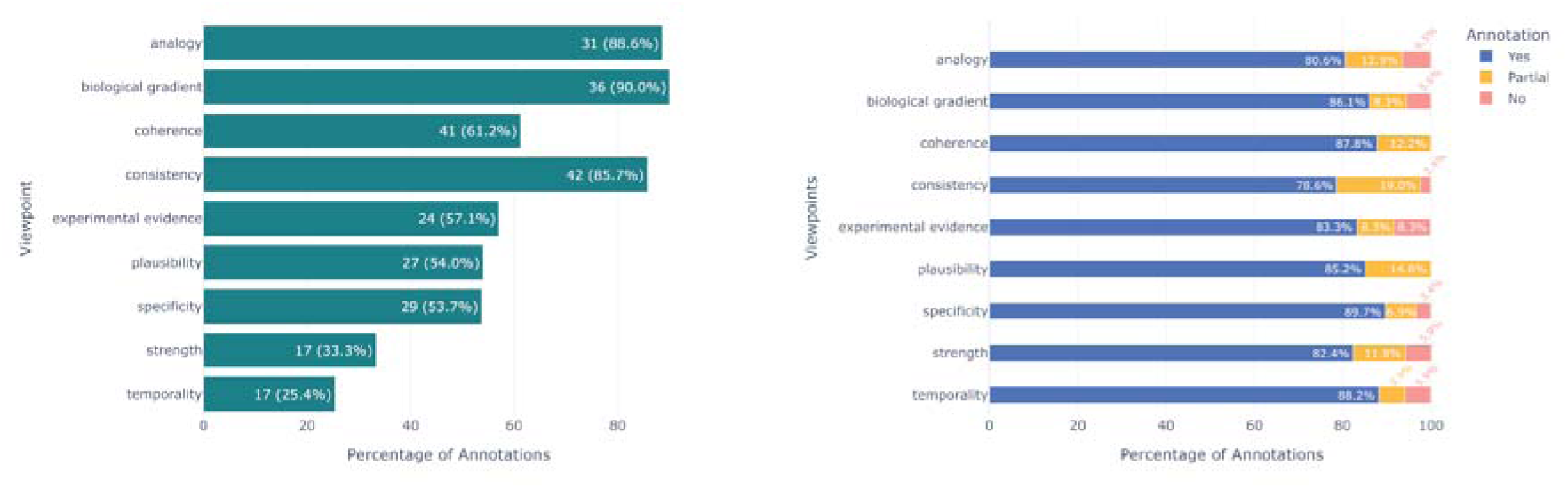
(left) Proportion of annotated sentences per viewpoint regardless of drug – adverse event combination. (right) Proportion of annotated sentences per viewpoint where the annotator agreed (yes), partially agreed (partial), and did not agree (no) with the output of the LLM.

The pattern for how well the annotator agreed with the output from the LLM is fairly consistent between the viewpoints, where the annotators agreed with the LLM on 77% - 90% of the output sentences. For sentences where the annotators partially agreed or disagreed, the proportions range from 6% - 19% and 0% - 8%, where *coherence* and *plausibility* were the viewpoints with no sentences the annotators fully disagreed with. See Figure 2 (right).

One pattern the annotators observed is that the LLM seems to favor de- and re-challenge as these aspects are mentioned in the output for several of the viewpoints while only being referred to in the viewpoint description of *strength, temporality* and *experimental evidence* given to the LLM. While information from the drugs’ summary of product characteristics (SPC) was not provided to the LLM via the prompts, it probably has inherent knowledge of them since they were likely part of the LLM’s vast training data. In one case the LLM incorrectly reasoned that since the adverse event of interest was not listed in the SPC of the drug, it is evidence against the association.

For the *strength* viewpoint, the LLM reasoned in a balanced way around the disproportionality measures provided without overstating their importance. However, it did not comment on their suitability for being used as a proxy for the effect size in a proper pharmacoepidemiological study.

For the viewpoints of *plausibility* and *coherence*, the LLM referred to specific reports multiple times even though these two viewpoints should not generally be reliant on the specific reports of the case series, but the combination of the specific drug and adverse event itself.

#### 3.2.2. Recall assessment

In the original *omeprazole – cutaneous vasculitis* signal text, 23 text segments were found to relate to, and subsequently mapped to, the BH viewpoints. All segments were mapped, but the viewpoints *coherence*, *biological gradient* and *experimental evidence* were not represented in the original signal text. The segments were also mapped to the LLM generated BH summaries. The signal text often reflects aggregated numbers and then drills downs to finer granularity. This aggregation is not seen in the generated LLM summaries, which makes it difficult to exactly match texts in the signal document and the LLM summaries. However, out of the 23 segments, 15 could be mapped to similar topic statements in the generated BH summaries. See Table 3.

**Table 3.** Number of segments per Bradford Hill viewpoint from the original signal text for omeprazole – cutaneous vasculitis that could be identified and mapped to the LLM output.

| Viewpoint | Segment count in original signal | Number of segments mapped to the LLM output |
| --- | --- | --- |
| Strength | 5 | 3 |
| Consistency | 3 | 1 |
| Specificity | 8 | 4 |
| Temporality | 3 | 3 |
| Biological gradient | 0 | 0 |
| Plausibility | 1 | 1 |
| Coherence | 0 | 0 |
| Experimental evidence | 0 | 0 |
| Analogy | 3 | 3 |
| **Total** | **23** | **15** |

One example where there was a match is for the *specificity* viewpoint, where the LLM successfully picked up on the importance of the histological findings that support the diagnosis of cutaneous vasculitis. See Table 4.

**Table 4.**
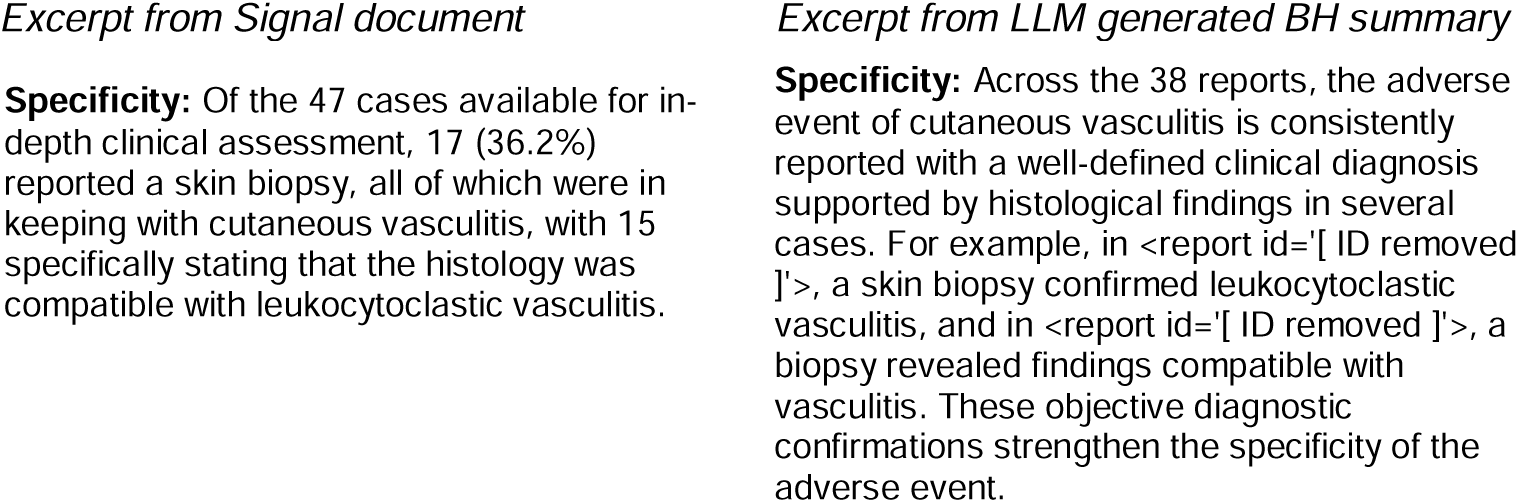
Example of where the LLM successfully picked up the importance of histological findings supporting the specificity viewpoint. Note that the difference in overall report count is due to reports updated after the original signal assessment were excluded from the study.

Eight of 23 signal segments did not match any LLM BH summary, four of those were related to the *specificity* viewpoint mentioning omeprazole as the sole suspected drug. However, the LLM touches on the topic in the summaries by mentioning “making it difficult to isolate omeprazole as the sole causative agent”. The signal segment “The earliest report was received in 1994 and the most recent in 2023.” related to *consistency* could not be mapped to a generated BH summary since report date was not fed to the LLM.

### 3.3. Manual review of overall assessments

The final summary output from the LLM for *omeprazole – cutaneous vasculitis* and the negative control *pazopanib – back pain* was manually reviewed for accuracy and is presented in Table 5. While there are areas that the LLM missed (as described in the previous section), the summaries highlight many important points to consider and present a balanced view of what the LLM found in its assessment.

**Table 5.**
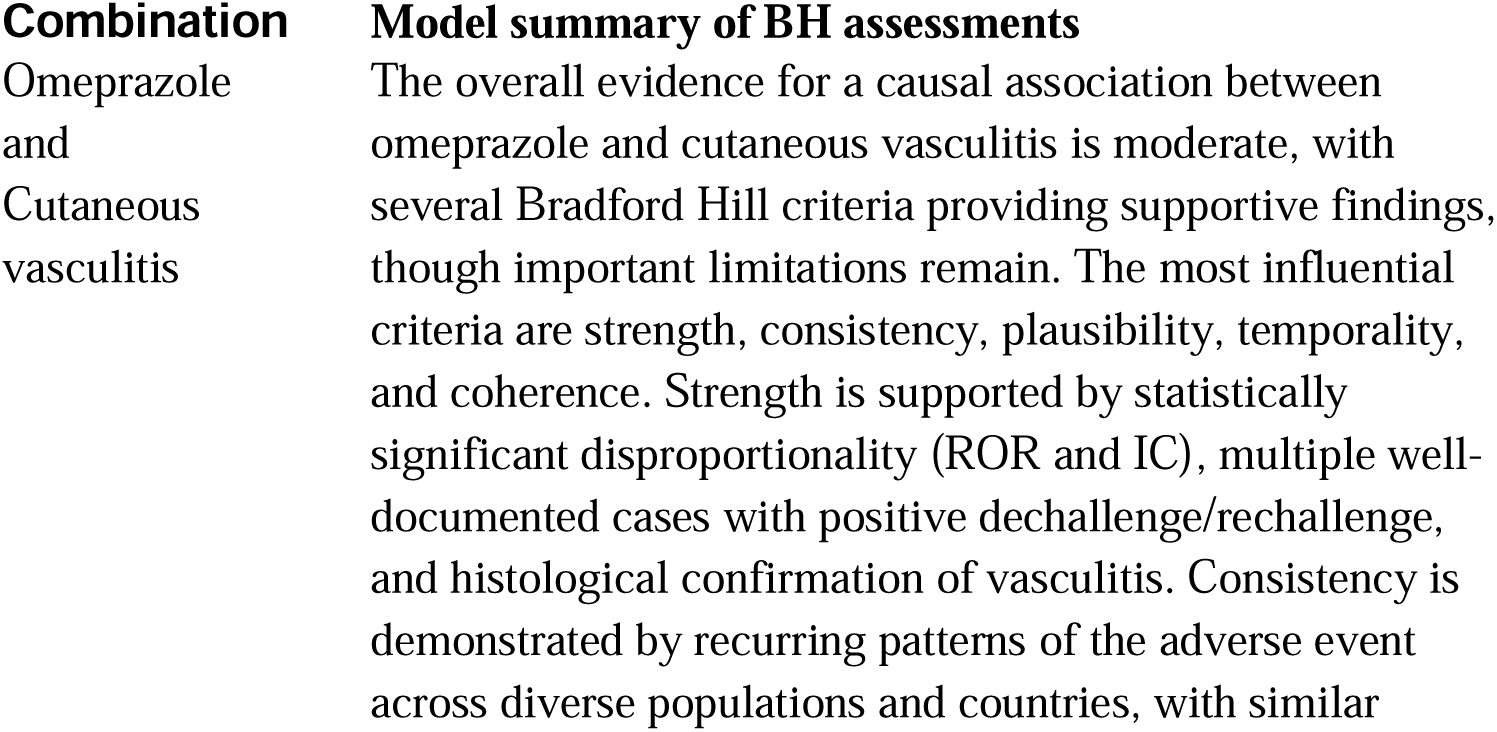
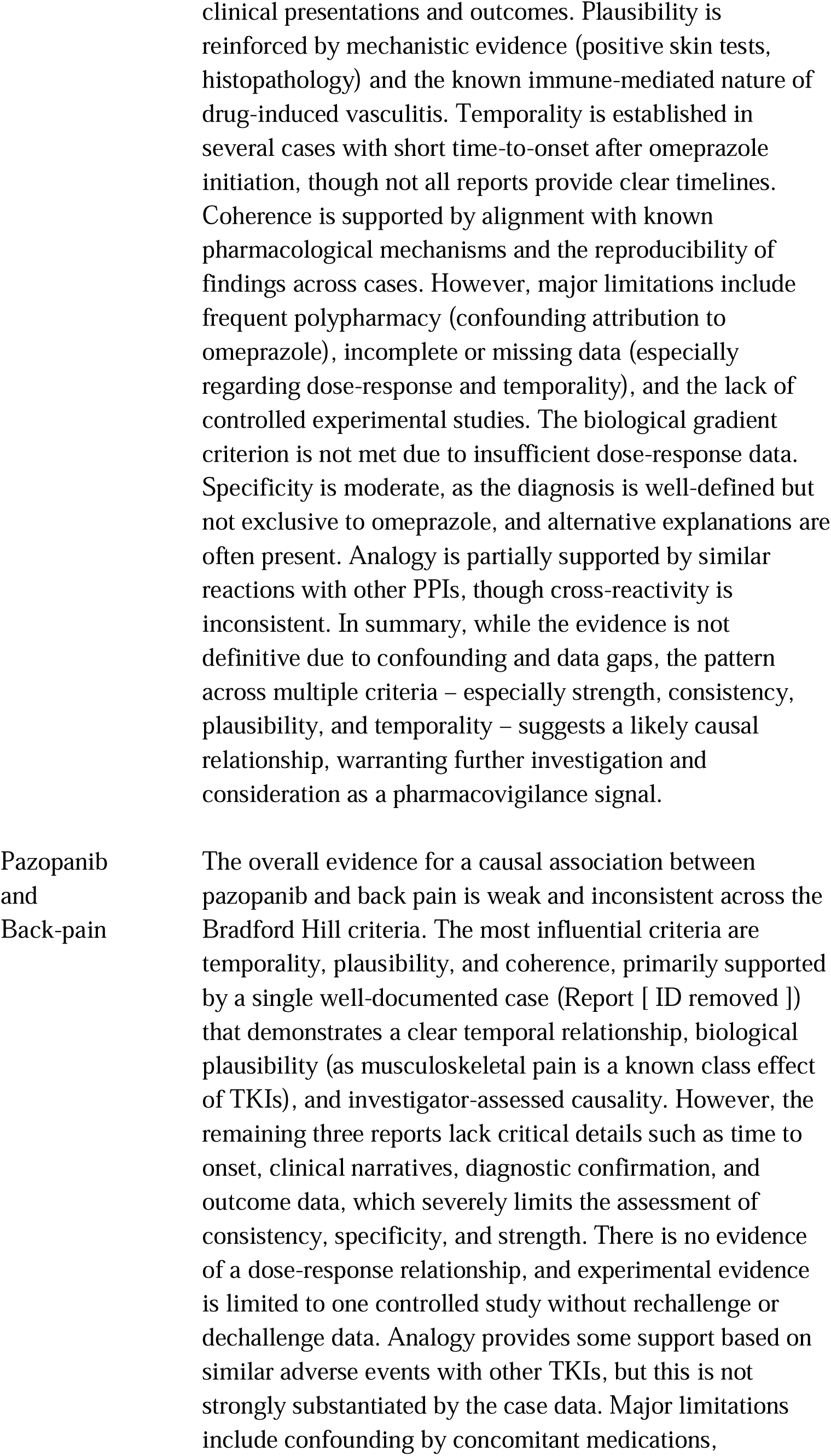
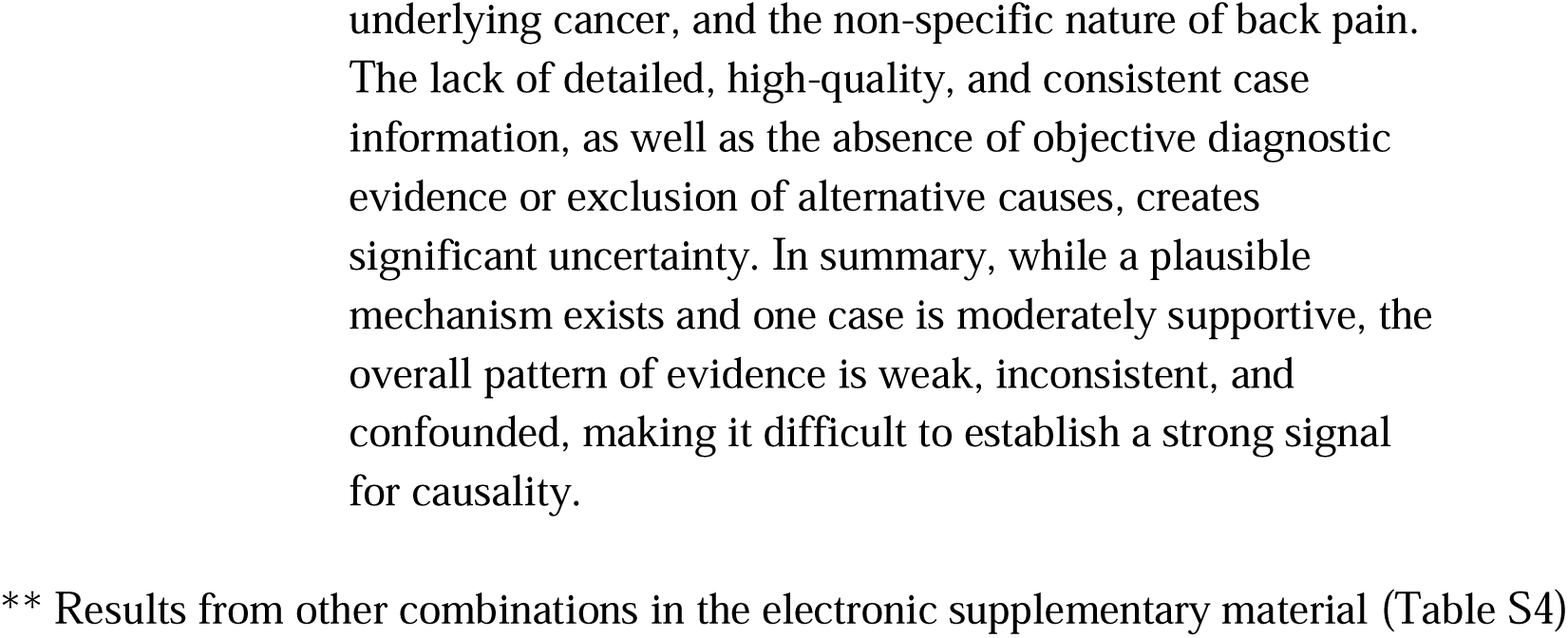
Overall summary output of the LLM’s causality assessment based on the LLM BH summaries.

## 4. Discussion

Our evaluation of how large-scale general-purpose LLMs can support PV professionals in assessing causality in a case series using BH viewpoints revealed three major insights: 1) that hallucinations were limited across all BH viewpoints and generally not severe, 2) that the LLM could produce comparable statements as made by humans but they were very dependent on the prompting, 3) that the final overall summaries were clear and balanced and can provide a useful starting point for the human assessors.

Our findings broadly align with current evidence on LLMs in PV: limited hallucinations are consistent with low rates reported in clinical evaluation frameworks [24], and the strong prompt-dependence we observed echoes findings across PV and healthcare LLM applications [25,26], underscoring the need for structured prompts in regulatory contexts. To our knowledge, this is the first study to apply LLMs to causality assessment at the case series signal assessment level, extending prior work on off-the-shelf LLMs for individual ICSR causality assessment [7] to a more complex and less well-defined task.

Our findings might have several implications for PV practice. This study focuses on the causality assessment step — the structured evaluation of a pre-identified drug – adverse event combination against a case series — rather than signal detection. Within this scope, we see potential for LLM-assisted assessment at two points in the workflow. Most immediately, once a validated signal requires in-depth assessment, an LLM can generate a structured preliminary review that the assessor can use as a starting point — valuable even for smaller case series, as it condenses case-level information, highlights patterns or angles worth investigating, and can surface early evidence against a signal. Further upstream, assessments could run in the background across a full prioritized list of drug – adverse event combinations before any human review, or on demand when an assessor picks up a case series. In all scenarios, we firmly position this as a support for human decision-making rather than a replacement for expert judgement.

In our study, the LLM provided a balanced interpretation of disproportionality analysis metrics and their statistical significance when assessing the BH *strength* viewpoint. The model appropriately considered the quantitative signals and contextualized them within the available case series data. However, it did not explicitly address the fundamental limitation that disproportionality analysis effect sizes are not suitable for determining the true strength of association as intended by the BH framework. However, we believe that improved prompting that explicitly informs the LLM of this would have produced better results. There might be value in the inclusion of disproportionality analysis metrics if more sub-analysis, such as stratification on patient age and sex, is provided along with prompting of common pitfalls of disproportionality analysis [27].

There were aspects from the original *omeprazole – cutaneous vasculitis* signal that were not covered in the LLM output. In these cases, there were aspects where we would not expect an LLM to perform well, such as being able to aggregate data on its own. Since the primary task of an LLM is to predict the next word in a sequence, they can be surprisingly bad at simple arithmetic operations, even though they are becoming better [28]. Here it would be helpful to pre-calculate such information and supply it to the LLM via the prompts.

The final overall summaries provided based on the nine separate per-viewpoint assessments show an impressive level of balanced reasoning that would be useful as a starting point for a human assessor. Especially considering it could be generated in a few minutes instead of days or weeks. Worth noting is that even though there were mistakes by the LLM in the per-viewpoint assessments, these did not result in an incorrect overall summary.

While these findings provide valuable insights into the potential and challenges of LLMs for causality assessment, several limitations should be acknowledged. First, the scope of our evaluation was qualitative in nature, and the data size was limited with only four positive and one negative control signals. While of course the number of statements assessed was larger, this limited scope may affect the generalizability of the results. Another limitation with regards to the data set worth highlighting is that our negative control (*pazopanib – back pain*) had a relatively low report count compared to the positive controls. Future studies should include multiple negative controls with report counts comparable to those of the positive controls. Similarly, it should be noted that the number of case reports available to the LLM was slightly lower than those available to the human assessors due to data extraction differences, e.g., 38 versus 47 reports for *omeprazole – cutaneous vasculitis*. For *dolutegravir – sexual dysfunction*, the LLM only had access to the five reports associated with the MedDRA PT term “sexual dysfunction” while the signals published as an abstract [19] assessed 348 reports after expanding the search to other relevant terms. Future work should consider enabling the LLM to perform such case series expansion, but it was not considered in this study since the scope of such a feature would have been too large.

Second, there exist limitations with regards to the prompts. The LLM did not have access to concomitant medications apart from co-suspected ones, making it impossible for it to make note of any non-suspected drugs that could be a possible confounder for the adverse event. Future prompt developments should carefully consider which data fields to include. Furthermore, having the prompt template include the case series data for all BH viewpoints caused the LLM to reason around the included reports, even for the viewpoints *plausibility* and *coherence* where this information might not be relevant. For these viewpoints, omitting the case series report data from the prompt might produce even better outputs.

Third, the *omeprazole – cutaneous vasculitis* combination was used both for developing the prompt template and for evaluating results. This was a pragmatic choice given the exploratory nature of this study, and we recognize that, as a general principle, prompt development and evaluation data should be kept separate. We note, however, that prompts are general natural language instructions rather than fitted parameters, and that the consistent performance across all other combinations — including the correct identification of the negative control — supports generalizability. Nevertheless, future studies should treat prompt development and evaluation cohorts as distinct.

A design choice worth addressing is the decision not to incorporate retrieval-augmented generation (RAG), a technique that supplements LLM prompts with externally retrieved information to improve grounding. We excluded it for two reasons: the exploratory nature of the study required keeping complexity manageable, and there is no obvious ready-made retrieval corpus for this task — the relevant knowledge spans pharmacological, epidemiological, and regulatory domains that are not easily packaged for retrieval. We therefore see RAG as a promising future direction rather than a missing component.

Additionally, models are rapidly evolving, and new state-of-the-art models are quickly emerging, with potential to show even more capabilities for this task in the future. In the same way, best practices for evaluating LLM-generated outputs are still developing for settings where the output space is unstructured text rather than a finite set of predefined categories. We opted for a precision and recall method, although other options are available. For example, using an LLM-as-a-judge [29] involves prompting a language model to evaluate results, allowing for large-scale assessments without much manual annotation. Automated semantic similarity metrics can also be used to compare outputs with gold standards, which is particularly helpful where ambiguity is minimal, a situation that doesn’t apply to signal assessment due to its variability and lack of standardization. Since we view the LLM’s assessment similarly to a human’s, in the sense that it would not produce the exact same output every time, we did not perform consistency checks between multiple runs. Doing this would have required an unfeasible amount of manual annotation. These considerations underscore the need for better evaluation frameworks for generative clinical outputs.

Future work should evaluate LLM performance on a larger and more diverse signal set, and assess how humans assisted by an LLM compare to either alone. These efforts would also benefit from better frameworks for evaluating generative output. A key finding of this study was the dependence on prompts to specify the task well, which aligns with previous studies [25,26]. However, while there exist quite specific methodologies for systematic case causality assessment [11,12], case series assessment is not well specified or harmonized in the PV field. We relied on BH-viewpoint instructions developed in this study, but our experience with human assessments is that assessors do not always follow a fixed protocol. Both human assessors and future automated approaches would benefit from clearer guidelines for performing and reporting case series signal assessments, in line with previously noted inconsistency in reporting of signals [30]. Several concrete methodological improvements are also worth exploring. As mentioned briefly earlier, pre-computing relevant summary statistics — such as time-to-onset, counts of single-suspected reports, and de- and rechallenge data — and providing these directly in the prompt would remove the need for the LLM to perform arithmetic on raw case data. Additionally, iterative self-critique approaches [31,32], where the LLM is prompted to act as a sceptic and critically evaluate and refine its own prior output, have been shown to improve output quality in summarization tasks [33,34]. Applying such an approach to the signal assessment workflow — potentially over several rounds — could yield more robust and balanced outputs.

This study raises the question whether generative models such as LLMs are ready for routine use in PV. Before being able to deploy them into production, regulatory compliance would be necessary including legal requirements such as the EU AI Act [35], as well as relevant guideline documents such as the CIOMS XIV report [36]. We argue for deploying such tools as support for human decision-making rather than full automation. However, they could even be considered as scoring mechanisms when prioritizing case series for manual assessment, assuming that the performance has been more thoroughly evaluated. A practical consideration is model lifecycle: off-the-shelf models accessed through a vendor may be retired and replaced, though newer versions are likely to represent capability improvements. With outputs always reviewed by a human assessor, a natural safeguard exists against undetected quality changes — but organizations should nonetheless establish a version monitoring and benchmarking protocol before transitioning to a new model in a production setting. Data privacy and security are also a prerequisite: sensitive case data cannot be processed through publicly accessible consumer interfaces, such as ChatGPT, Claude, or Copilot, but instead requires a controlled environment with appropriate safeguards, such as the private Azure OpenAI deployment used in this study (Section 2.2).

## 5. Conclusion

This study demonstrates that LLMs can provide balanced and contextually nuanced support for causality assessment in PV, particularly when carefully prompted to perform signal assessments of case series in line with the BH viewpoints.

The LLM was able to synthesize complex case information and offer reasoned summaries that align with human expert assessments, while maintaining a low rate of hallucinations and overstatements. However, our findings also highlight important limitations. Even though the rate of hallucinations was low, it was not zero, and it is difficult to predict how they would affect the final output. This underscores the need for careful human oversight and methodological rigor when integrating LLMs into PV workflows. As GenAI continues to evolve, future research should focus on refining prompt strategies, improving the interpretability of model outputs, and developing robust frameworks for evaluating AI-assisted signal assessment. Ultimately, LLMs hold promise as valuable tools to augment, but not replace, expert judgment in drug safety evaluation.

## Data Availability

Access to the case series data is restricted based on the data access conditions for access to data for the WHO Global Databases of adverse events of medicines and vaccines. Subject to these conditions, data is available from the authors on reasonable request.

## 6. Declarations

## Acknowledgements

We would like to acknowledge Carlos Melgarejo-González contributions during early phases of this study, and Daniele Sartori, G Niklas Norén, Deliana Aboka and Qun-Ying Yue for reviewing the manuscript. Co-author Joseph Mitchell, as of August 2025, is no longer employed by UMC. However, their contributions to the study were made prior to the time of departure, as part of their employment at UMC.

The authors are indebted to the members of the WHO Programme for International Drug Monitoring who contribute reports to VigiBase. However, the opinions and conclusions of this study are not necessarily those of the various member organisations nor of the WHO.

MedDRA^®^ trademark is registered by ICH.

## Ethics approval and consent to participate

Not applicable. The data in VigiBase is de-identified and not considered personal data.

## Competing interests

The authors declare that they have no competing interests.

## Author contributions

AS, AZ, AV, ELM and HTG conceptualized and designed the study with support from the other co-authors (JM, JB and LS). The method and study design were informed by a prelJstudy conducted by JB and LS with support from ELM. AS, AV and HTG conducted the LLM runs. AS, AV, AZ and HTG performed annotations. AS and HTG wrote the manuscript with support from all co-authors, who also reviewed and approved drafts and the final version.

## Footnotes

^1^Generally accessible in the sense that it is not a limited release, so anyone could access it without filling out an application or waiting in a queue.

^2^MedDRA^®^ the Medical Dictionary for Regulatory Activities terminology is the international medical terminology developed under the auspices of the International Council for Harmonisation of Technical Requirements for Pharmaceuticals for Human Use (ICH). It consists of over 80,000 terms organized into a hierarchy of five levels. System Organ Class (SOC, e.g., gastrointestinal disorders), High Level Group Term (HLGT, e.g., gastrointestinal signs and symptoms), High Level Term (HLT, e.g., nausea and vomiting symptoms), Preferred Term (PT, e.g., nausea), and Lowest Level Term (LLT, e.g., feeling queasy).

## Notes

### Competing Interest Statement

The authors have declared no competing interest.

### Author Declarations

Ethical board review was not required for this study, as this study did not use personal data. Data was used according to the Data Access Conditions from VigiBase, the WHO Global Databases of adverse events of medicines and vaccines.

