## Supplementary material for "AI as a signal assessor – Can a Large Language Model perform causality assessment on a case series?"

Electronic Supplementary Material

Journal: **Drug Safety**

Main article title:

### Section 1: Bradford Hill (BH) viewpoint descriptions

Below are the BH viewpoint descriptions fed to the LLM for this study.

#### *Strength*

```
Strength of association is a Bradford-Hill criterion that reflects how strongly the
observed data support a relationship between a drug and an adverse event.

This involves comparing the number of reported cases (observed) to the number
expected in the absence of a causal association. In this case:

- **Total reports in database**: {{ disprop_details.n_total | int }}
- **Reports involving the drug**: {{ disprop_details.n_drug | int }}
- **Reports involving the reaction**: {{ disprop_details.n_reaction | int }}
- **Observed number of cases**: {{ disprop_details.n_observed }}
- **Expected number of cases**: {{ disprop_details.n_expected | round(2) }}

A higher ratio of observed to expected cases in VigiBase—the global database of
individual case safety reports—may indicate a stronger potential signal.

Disproportionality analyses are used to assess this:

- **Reporting Odds Ratio (ROR)**: {{ disprop_details.ror | round(2) }} (95% CI
lower bound: {{ disprop_details.ror025 | round(2) }})
- **Information Component (IC)**: {{ disprop_details.ic | round(2) }} (95% CI lower
bound: {{ disprop_details.ic025 | round(2) }})

In addition to statistical measures, qualitative strength of association is also
considered. This involves evaluating cases that provide compelling clinical
evidence for causality, such as those with a positive dechallenge (where the event
resolves after drug withdrawal) or a positive rechallenge (where the event recurs
after re-administration). The presence of multiple such cases especially using only
dechallenge or rechallenge information can further strengthen the association.
```

#### *Consistency*

```
Consistency, also known as reproducibility, is a Bradford Hill criterion that
refers to the extent to which an observed association between a drug and an adverse
event is consistently seen across different settings, populations, and time
periods. In pharmacovigilance, this criterion is supported when similar drug-event
combinations are reported across multiple independent sources (e.g., different
countries, healthcare systems, or reporters); over time, suggesting a stable and
non-random signal; in diverse populations (e.g., age groups, sexes, comorbidities),
and with similar clinical manifestations and outcomes (e.g., time to onset,
severity, reversibility). Consistency strengthens causal inference by reducing the
likelihood that the observed association is due to chance, bias, or confounding.
The presence of aligned findings across spontaneous reporting systems, published
literature, or observational studies adds further weight to this criterion.
```

#### *Specificity*

```
Specificity is a Bradford Hill criterion that evaluates the extent to which the
association between a drug and an adverse event is limited to a well-defined
diagnosis and a narrow set of circumstances, thereby reducing alternative
explanations. In pharmacovigilance case series, this includes assessing how
clinically specific the reported outcome is—for example, "appendicitis" is more
specific than "abdominal pain"—and whether the diagnosis is supported by objective
evidence such as biopsy results, laboratory findings, or diagnostic tests.
```

Causation is considered more likely when the event occurs under a very specific set of circumstances. Specificity also involves evaluating alternative hypotheses for the adverse event, such as underlying diseases, patient risk factors, or concomitant medications. High specificity strengthens causal inference when the event is rare, well-characterized, and unlikely to be explained by other factors.

#### *Temporality*

Temporality is a Bradford Hill criterion that assesses whether the timing of an adverse event is compatible with a causal relationship to the suspected drug. For a causal inference to be plausible, the adverse event must occur after the administration of the drug. This criterion also considers whether the time to onset (TTO) is consistent with what is known about the pharmacokinetics and pharmacodynamics of the drug (e.g., absorption, distribution, metabolism, elimination), the pathophysiological mechanism underlying the adverse event, and established clinical patterns or previously reported case series. When evaluating temporality in a case series, it is important to consider whether the adverse event occurred after drug exposure, whether the time interval between exposure and onset is biologically plausible, and whether there is supportive dechallenge (i.e., improvement after stopping the drug) or rechallenge (i.e., recurrence upon re-exposure) information.

#### *Biological gradient*

Biological Gradient, also known as the dose-response relationship, is a Bradford Hill criterion that evaluates whether changes in the dose or exposure level of a drug are associated with corresponding changes in the occurrence or severity of an adverse event. A positive gradient is typically observed when higher doses lead to more frequent or more severe adverse events, supporting a causal link. However, inverse or non-linear relationships may also be relevant, as long as a consistent and biologically plausible pattern is observed. Assessing this criterion involves examining whether reported cases show a trend between dose variations and adverse outcomes.

#### *Plausibility*

Plausibility is a Bradford Hill criterion that refers to whether a biologically or pharmacologically credible mechanism exists to explain how the drug could cause the reported adverse event, based on current scientific understanding. This includes known pharmacodynamics, pharmacokinetics, drug class effects, and relevant pathophysiological mechanisms. The focus is on the existence of a scientifically reasonable basis for causation, rather than on individual patient-specific factors. While the primary focus is on mechanistic plausibility at the general level, case-specific factors (e.g., patient characteristics, concomitant medications or underlying diseases) could also have an impact on the biological plausibility of the reaction.

#### *Coherence*

Coherence is a Bradford Hill criterion that assesses whether the observed association between a drug and an adverse event aligns with existing scientific knowledge. This includes the drug’s known pharmacological properties, mechanism of action, pharmacokinetics, and previously documented safety profile. Coherence also considers whether the event aligns with what is known about the condition, integrating epidemiological, clinical, and biological context — including known adverse effects, patient characteristics, and the timing and route of drug administration. Additionally, coherence accounts for whether the event makes sense in the patient’s specific context — for example, an adverse event rooted in a congenital or genetically determined condition is unlikely to be caused by a drug first administered in adulthood. In summary: Given the known properties of the drug and existing scientific knowledge, does it make clinical and biological sense that the drug could have caused this reaction?

#### *Experimental evidence*

Experimental evidence is a Bradford–Hill criterion relating to whether data derived from controlled investigations that test hypotheses about causal relationships exists. This typically includes findings from randomized controlled trials (RCTs), biological or mechanistic studies, and animal models that explore the effect of the drug on physiological systems. Such evidence strengthens causal inference by demonstrating reproducibility under controlled conditions. Supportive evidence may also come from longitudinal observational studies or from individual case report features such as positive dechallenge (symptom resolution after drug withdrawal) or rechallenge (symptom recurrence upon reintroduction); however, these latter forms are generally considered weaker and less conclusive compared to formal experimental data.

#### *Analogy*

Analogy is a Bradford–Hill criterion that evaluates whether similar drugs (e.g., from the same pharmacological class or with similar mechanisms of action) have been associated with the same or comparable adverse events. If such analogous associations have been observed—either with structurally related compounds or drugs with similar therapeutic profiles, this precedent may strengthen the case for causality in the drug currently under investigation. The presence of analogies lends weight to causality, especially when supported by clinical or mechanistic rationale. However, the absence of analogous associations or a mechanistic explanation does not rule out the possibility of causality, as unique properties of the drug or context-specific factors may still contribute to the event. Absence of analogy does not negate a causal link; it may simply reflect limited data for newer or less-studied drugs.

### Section 2: LLM Prompts

**Note: The prompts in this supplement include instructions of generating a score indicating the strength of the evidence. However, these scores were not used or analyzed in this study.**

**Note: In the prompts, each Bradford Hill viewpoint description is labeled a "definition." In the main text and Section 1 above we instead use "description" to avoid implying an authoritative interpretation of the Bradford Hill viewpoints.**

Table S1: Optimized prompt used for experiments

**System prompt:**

```
You are an experienced medical doctor and clinical pharmacist working as a
pharmacovigilance assessor at a national pharmacovigilance centre. Your task is
to evaluate individual case safety reports (ICSRs) to assess the likelihood that
a reported adverse event was caused by a specific medicine.

You are currently reviewing a case series involving:
- **Suspected Drug**: {{ drug name }}
- **Adverse Event**: {{ adverse event }}
- **Number of Cases**: {{ number_of_cases }}
```

Each ICSR may include:
- Patient demographics
- Details of the suspected medicine(s), including dose and duration
- Description of the adverse event, including time to onset
- A narrative of the clinical course, including outcomes, dechallenge/rechallenge, and any relevant lab or diagnostic findings and medical history

Please note:
- These reports are **suspected** adverse reactions and may be submitted by healthcare professionals or patients.
- The information may be **incomplete or imprecise**. Do not assume missing details or invent information.
- If the data is insufficient to make a confident assessment, clearly state that.

Your goal is to:
1. **Assess causality** using clinical reasoning and pharmacological knowledge.
2. **Identify patterns** across the case series that may support or weaken a causal relationship.
3. **Apply relevant Bradford Hill criteria that is stated** (e.g., temporality, dechallenge/rechallenge, biological plausibility, consistency, specificity, dose-response).

Your assessment will inform human decision-making in pharmacovigilance. Accuracy and transparency are critical, as incorrect conclusions may impact patient safety.

**User prompt:**

You are assessing a case series of individual case safety reports of suspected side effects to medicines, using the Bradford-Hill criteria for causality. These are a set of 9 categories of evidence to support an assessment of whether or not a side effect could plausibly be caused by a medicine.

For this task, you will focus your assessment on **{{ bradford_hill_criterion }}**.

#### Criterion Definition ###
{{ bradford_hill_definition }}

#### CASE SERIES DATA ###
This case series consists of {{ n_reports }} individual reports. Below are the structured summaries:

<case_series>
{{ case_series_content }}
</case_series>

{% if focus != '' %}
#### Additional Focus ###
{{ focus }}
{% endif %}

```
### TASK ###
Analyze the above reports specifically with regard to the criterion of **{{
bradford_hill_criterion }}**. Look for common patterns, differences, and
strengths or weaknesses in the evidence.

If you cite specific cases, refer to them by their `<report id="...">`.

Do **not** reflect on whether this affects overall causality — focus only on the
selected criterion.

### OUTPUT FORMAT ###
Respond with valid JSON (not inside a code block or markdown) in this format:
{
"summary": "<Detailed synthesis of the criterion across the case series>",
"score": x
}
Where **x** is an integer from 0 (no evidence) to 5 (strong evidence).
```

**Example LLM Response:**

```
{"summary":

"The plausibility of omeprazole causing cutaneous vasculitis is supported
by several mechanistic and clinical observations across the case series,
but the evidence is not uniformly strong. Omeprazole is a proton pump
inhibitor (PPI) that can modulate immune responses, potentially leading
to hypersensitivity reactions, including vasculitis. Cutaneous vasculitis
is often mediated by immune complex deposition and neutrophilic
infiltration, which aligns with the histological findings (e.g.,
leukocytoclastic vasculitis) reported in several cases. Specific cases,
such as <report_id="[ ID removed ]"> and <report_id="[ ID removed ]">,
provide stronger mechanistic support due to biopsy-confirmed
leukocytoclastic vasculitis and positive skin patch tests implicating
omeprazole. Additionally, temporal relationships between omeprazole
initiation and symptom onset (e.g., <report_id="[ ID removed ]"> and
<report_id="[ ID removed ]">) strengthen the plausibility. However,
confounding factors, such as concomitant medications (e.g., NSAIDs,
antibiotics) and underlying conditions (e.g., autoimmune diseases,
infections), weaken the specificity of the association in many cases.
Furthermore, rechallenge data is limited, and in some cases, the reaction
resolved after discontinuation of multiple drugs, making it difficult to
isolate omeprazole as the causative agent. Overall, while there is a
biologically plausible mechanism and some supporting clinical evidence,
the presence of confounders and lack of consistent rechallenge data
reduce the strength of the evidence.",

"score": 3}
```

Minor prompt formatting changes, such as removing line breaks, did not affect the content. Additionally, we observed that it was useful to define the output format as below instead of ‘Respond with JSON’ to make sure the response from the model was not wrapped in a string <*′′′json ′′′*>.

```
### OUTPUT FORMAT ###
Respond with valid JSON (not inside a code block or markdown) in this format:
{
  "summary": "<Detailed synthesis of the criterion across the case series>",
  "score": x
}
Where **x** is an integer from 0 (no evidence) to 5 (strong evidence).
```

Structuring the prompt with headers like ‘TASK’ and ‘OUTPUT FORMAT’ helps direct the response into a more predictable format (Table S3). For example, asking the model to provide a summary in a valid JSON format allows for a concise overview, and the score provided by the model is a self-assessment of the evidence it has collected with respect to the combination.

Table S2: Effect of adding caution to the system prompt

**Response:**

```
### Assessment of Biological Plausibility: Omeprazole and Cutaneous
Vasculitis

#### Overview of Omeprazole
Omeprazole is a proton pump inhibitor (PPI) that irreversibly inhibits
the H+/K+ ATPase enzyme in gastric parietal cells, leading to suppression
of gastric acid secretion. It is widely used for conditions such as
gastroesophageal reflux disease (GERD), peptic ulcers, and Zollinger-
Ellison syndrome. Omeprazole is generally well-tolerated, but it has been
associated with a range of adverse effects, including hypersensitivity
reactions and rare immune-mediated conditions.

#### Overview of Cutaneous Vasculitis
Cutaneous vasculitis refers to inflammation of the small blood vessels in
the skin, often presenting as palpable purpura, erythematous macules, or
ulcers. It can be caused by infections, autoimmune diseases,
malignancies, or drugs. Drug-induced vasculitis is typically immune-
mediated and involves hypersensitivity mechanisms, often with deposition
of immune complexes in the vessel walls, leading to complement
activation, inflammation, and vascular damage.

#### Biological Plausibility of the Association
The potential link between omeprazole and cutaneous vasculitis can be
evaluated based on known pharmacological and immunological mechanisms:

1. **Immune-Mediated Hypersensitivity Reactions**:
   - Omeprazole has been implicated in hypersensitivity reactions,
including rare cases of systemic and cutaneous immune-mediated conditions
such as Stevens-Johnson syndrome, toxic epidermal necrolysis, and drug
rash with eosinophilia and systemic symptoms (DRESS). These reactions
suggest that omeprazole can act as a trigger for immune dysregulation.
   - Drug-induced vasculitis is often mediated by a type III
hypersensitivity reaction, where immune complexes (e.g., drug-antibody
complexes) deposit in small blood vessels, activating the complement
cascade and leading to inflammation and vessel damage. While direct
evidence of omeprazole forming immune complexes is lacking, its ability
to induce other immune-mediated conditions supports the plausibility of
this mechanism.

2. **Role of Reactive Metabolites**:
```

- Omeprazole is metabolized primarily in the liver by cytochrome P450 enzymes (CYP2C19 and CYP3A4). Although omeprazole is not typically associated with the formation of highly reactive metabolites, minor reactive intermediates could theoretically bind to proteins and form neoantigens, triggering an immune response. This mechanism has been proposed for other drugs associated with vasculitis.

3. **Drug Class Effects**:
   - Other PPIs, such as lansoprazole and pantoprazole, have also been associated with rare cases of vasculitis, suggesting a potential class effect. This supports the idea that PPIs, including omeprazole, may share a common mechanism that predisposes to immune-mediated vascular inflammation.

4. **Alteration of Immune Homeostasis**:
   - PPIs, including omeprazole, have been shown to modulate immune function. For example, they can inhibit lysosomal acidification in antigen-presenting cells, potentially altering antigen processing and presentation. This could theoretically contribute to immune dysregulation and hypersensitivity reactions, including vasculitis.

5. **Potential Role of Off-Target Effects**:
   - Omeprazole has been reported to have off-target effects, such as inhibition of carbonic anhydrase and modulation of endothelial function. While these effects are not directly linked to vasculitis, they could theoretically contribute to vascular or immune dysregulation in susceptible individuals.

6. **Systemic Inflammation and Cytokine Release**:
   - Drug-induced vasculitis often involves the release of pro-inflammatory cytokines (e.g., TNF-α, IL-6) and chemokines that recruit neutrophils and other immune cells to the vessel wall. While omeprazole itself does not directly induce cytokine release, its role in triggering immune dysregulation could indirectly lead to this inflammatory cascade.

##### Case-Specific Factors
While the general mechanisms above provide a plausible basis for the association, case-specific factors such as the patient’s underlying immune status, concomitant medications, or pre-existing autoimmune conditions could significantly influence the likelihood of omeprazole-induced vasculitis. For example:
- **Concomitant Medications**: Other drugs taken by the patient may contribute to immune complex formation or synergize with omeprazole to trigger vasculitis.
- **Genetic Predisposition**: Polymorphisms in drug-metabolizing enzymes (e.g., CYP2C19) or immune-related genes could increase susceptibility to hypersensitivity reactions.

##### Conclusion
The association between omeprazole and cutaneous vasculitis is biologically plausible based on the following:
- Known ability of omeprazole to trigger immune-mediated hypersensitivity reactions.
- Potential for immune complex formation and complement activation, leading to vascular inflammation.
- Evidence of similar adverse events with other PPIs, suggesting a possible class effect.
- Modulation of immune function by omeprazole, which could predispose to immune dysregulation.

While direct evidence linking omeprazole to cutaneous vasculitis is limited, the mechanistic pathways described above provide a scientifically reasonable basis for causation. Further research into the immunological effects of omeprazole and its metabolites would help clarify this association.

### Table S3: Effect of adding section headers in the user prompt

**Appended to user prompt:**

```
### TASK ###

Based on your knowledge of pharmacology, pathophysiology, and drug mechanisms, explain the biological plausibility of this association. Identify any molecular targets, pathways, pharmacokinetic or pharmacodynamic properties, or systems-level effects that could explain a causal link.

### OUTPUT FORMAT ###

Respond with valid JSON (not inside a code block or markdown) in this format: {

"summary": "", "score":

} where x is an integer from 0 (no evidence) to 5 (strong evidence).
```

**Response:**

```
{
  "summary": "Omeprazole, a proton pump inhibitor (PPI), works by irreversibly inhibiting the H+/K+ ATPase enzyme in gastric parietal cells, reducing gastric acid secretion. While its primary mechanism of action is unrelated to immune or vascular processes, there is biological plausibility for cutaneous vasculitis as an adverse event. PPIs, including omeprazole, have been associated with hypersensitivity reactions, likely mediated by immune mechanisms. Drug-induced vasculitis is often linked to immune complex deposition, complement activation, or direct drug effects on vascular endothelium. Omeprazole has been reported to trigger immune-mediated adverse events, such as drug-induced lupus erythematosus, which shares some pathophysiological features with vasculitis. Additionally, omeprazole metabolism involves the cytochrome P450 system (primarily CYP2C19 and CYP3A4), and reactive metabolites or oxidative stress could theoretically contribute to endothelial damage or immune activation. While the exact mechanism remains speculative, the immune-mediated and inflammatory pathways provide a biologically plausible basis for the association.",
  "score": 3
}
```

**Table S4: Overall summary of causality assessment using the BH summaries.**
**Important note: These summaries have not been manually verified for accuracy.**

| Combination | Model summary |
| --- | --- |
| Covid-19 vaccine and Vulval-ulceration | The evidence for a causal association between Covid-19 vaccines and vulval ulceration is supported by several influential Bradford Hill criteria. The strength of the association is notable, with a high reporting odds ratio (ROR 5.25) and a significant disproportionality signal, indicating that vulval ulceration is reported more frequently than expected after Covid-19 vaccination. Consistency is demonstrated by similar clinical presentations, temporal patterns (onset within 1-3 days), and recurrence after subsequent doses across diverse geographic and demographic groups. Temporality is robust, with most cases occurring shortly after vaccination, and some rechallenge cases further supporting a temporal link. Specificity is moderate, as many cases include negative diagnostic workups for alternative causes, though some reports lack detail or involve confounders. Plausibility and coherence are supported by the known immune-stimulating properties of vaccines and the alignment with immune-mediated mechanisms, especially in predisposed individuals. However, the evidence is weakened by the lack of direct mechanistic or controlled experimental studies, incomplete data in some reports, and the absence of a clear biological gradient (dose-response relationship). Analogy is partially supported by similar reactions across different Covid-19 vaccine platforms, but not beyond this class. Major limitations include confounding factors (e.g., pre-existing conditions), variable report quality, and the rarity of the event, which limits generalizability. Overall, the pattern of evidence is strongest for strength, consistency, temporality, and plausibility, but is tempered by gaps in experimental and mechanistic data and some inconsistencies in reporting. |
| Dolutegravir and Sexual dysfunction | The evidence for a causal association between Dolutegravir and sexual dysfunction is weak and inconsistent across the Bradford Hill criteria. The most influential criteria – consistency, temporality, and strength – show repeated reports of sexual dysfunction in diverse settings, with some cases occurring shortly after drug initiation and one case demonstrating resolution after withdrawal (positive dechallenge). However, these signals are undermined by major limitations: small case numbers, lack of detailed clinical narratives, broad and poorly defined adverse event descriptions, and the presence of multiple concomitant medications that confound attribution. The strength of association is statistically weak (low ROR and IC, with confidence intervals crossing null), and |

specificity is poor due to alternative explanations and lack of objective diagnostic evidence. Plausibility and coherence are limited by the absence of a direct pharmacological mechanism and inconsistent alignment with Dolutegravir's known safety profile. No dose-response relationship is evident, and experimental evidence is restricted to a single dechallenge case. Analogy is weak, as similar drugs do not have well-established links to sexual dysfunction. Overall, while a non-random pattern is suggested, the quantity and quality of evidence are insufficient to support a strong or specific causal signal, and uncertainty remains high due to confounding and data gaps.

| | |
|---|---|
| Methotrexate and Muscle-spasms | The overall evidence for a causal association between methotrexate and muscle spasms is weak to moderate, with several important limitations. The most influential criteria are consistency, temporality, and plausibility. Consistency is moderately supported by multiple case reports from diverse demographics and settings, showing muscle spasms occurring with methotrexate use, sometimes resolving after withdrawal. Temporality is also moderately supported, with several cases documenting symptom onset after methotrexate initiation and improvement upon discontinuation. Plausibility is present, as methotrexate could theoretically cause muscle spasms via mechanisms such as electrolyte disturbances, but this is not strongly established. However, the strength of association is low: disproportionality analyses (ROR, IC) show no signal in the broader database, and the number of observed cases is lower than expected. Specificity is weak due to frequent confounding by comorbidities and concomitant medications, and the clinical presentations are variable. There is no clear biological gradient (dose-response), and experimental evidence is limited to a few dechallenge/rechallenge cases without controlled studies. Coherence is limited by the absence of muscle spasms as a recognized adverse effect in methotrexate's established safety profile. Analogy is only moderately supported, as similar events are reported with other drugs but with similar confounding. Major gaps include lack of controlled studies, inconsistent dose-response, and frequent confounding factors. In summary, while there are some case-level signals and plausible mechanisms, the overall pattern is inconsistent and not robust, with significant uncertainty and alternative explanations. |
